# Hand Hygiene Practices in Paediatric Populations: Assessing Their Impact on Infectious Disease Outbreaks in Preschools and Schools. [Protocol]

**DOI:** 10.1101/2025.07.02.25330765

**Authors:** Alan Silburn Grad, Namita Singh

## Abstract

Infectious diseases remain a significant concern in educational settings, particularly among preschool and school-aged children, who are vulnerable to respiratory and gastrointestinal illnesses due to underdeveloped immune systems and frequent interpersonal interactions (1). Hand hygiene has proven effective in preventing infections in healthcare settings (2); however, its impact on educational environments has not been thoroughly explored. This protocol outlines a systematic review to assess the relationship between hand hygiene practices and the prevalence of paediatric infectious disease outbreaks in schools and preschools. By synthesising evidence from studies on the effectiveness of hand hygiene interventions, the review will aim to identify key moments for intervention and explore how structured hygiene programs could be adapted from hospital settings to reduce disease transmission. The findings are expected to inform public health strategies and policy development, guiding the implementation of hand hygiene protocols to improve health outcomes in educational environments.

## 1. Background

### I. Important characteristics

Maintaining proper hand hygiene is a fundamental preventive measure in public health, particularly in settings with vulnerable populations, such as young children. Children are particularly vulnerable to infectious diseases due to their underdeveloped immune systems, frequent interpersonal interactions, and shared resources in settings like preschools and schools (1). These environments may contribute to the rapid spread of respiratory infections (e.g., influenza, common cold, respiratory syncytial virus) and gastrointestinal illnesses (e.g., norovirus, salmonella, campylobacter, Escherichia coli). Inadequate hand hygiene practices among staff and students may contribute to the rapid spread of pathogens, resulting in increased rates of communicable infections. These outbreaks not only pose serious health risks but also disrupt educational activities, affecting both learning outcomes and the well-being of children (3).

### II. Literature Review

Maintaining proper hand hygiene is a fundamental preventive measure in public health, particularly in settings with vulnerable populations, such as young children. Inadequate hand hygiene practices among staff and students contribute significantly to the rapid spread of pathogens, resulting in increased rates of respiratory and gastrointestinal infections. These outbreaks not only pose serious health risks but also disrupt educational activities, affecting both learning outcomes and the well-being of children (3).

The success of the Five Moments for Hand Hygiene program in hospital settings demonstrates the transformative impact of structured hand hygiene initiatives on reducing infection rates. By emphasising key moments for hand hygiene: before patient contact, before aseptic tasks, after exposure to body fluids, after patient contact, and after contact with patient surroundings, this program has significantly reduced health-care-associated infections worldwide (4). Its effectiveness inspires a model that could be adapted for school environments to improve hand hygiene practices and reduce infectious disease transmission. Implementing a similar structured approach to hand hygiene in preschools, focusing on critical moments such as before handling food, after using the restroom, and after playing with shared toys, could help mitigate the occurrence of respiratory outbreaks and promote overall health and well-being among this population. However, at present, no such initiative is as universally adopted.

This literature review aims to assess the impact of hand hygiene practices on infectious disease outbreaks in preschool settings, with a focus on paediatric populations, acting as a foundation for the greater systematic review project proposed. Ultimately, the project will provide valuable insights to guide the development and implementation of effective hand hygiene interventions tailored for different educational environments.

Paediatric infectious disease outbreaks are a significant concern for the public, as they impact school attendance and parental productivity, with educational and economic implications. These outbreaks persist in younger age groups, particularly in school and preschool-aged children. While interventions like contact precautions, vaccinations, school closures, and isolation can help mitigate disease transmission, the relationship between children’s hand hygiene practices is less well-documented.

This review highlights the essential role of hand hygiene practices in mitigating the transmission of infectious diseases in preschool and community settings. Torner et al. (5) demonstrated that frequent handwashing significantly reduces the risk of paediatric influenza, particularly in school-aged children, underscoring its importance as a protective factor against infectious disease outbreaks. This finding aligns with the conclusions of the Hi Five study (6), which conducted a school-based randomised trial in Denmark, revealing that improved hygiene practices effectively reduce childhood infections. Importantly, the Hi Five study emphasises the broader social implications of infectious illnesses, including school absenteeism, disruptions to parental work, and increased public healthcare costs. These findings synergistically support the World Health Organization’s determination that handwashing is a cornerstone of primary prevention (4).

The COVID-19 pandemic has further underscored the necessity of stringent hygiene practices. Matsuda et al. (7) examined behavioural changes in children’s preventive activities during the pandemic, highlighting the opportunity for long-term improvements in hygiene behaviours. However, despite the evidence supporting hand hygiene as a key intervention, further research is essential to address its specific impact in preschool environments. This includes evaluating the feasibility and sustainability of implementing hand hygiene programs in diverse settings and understanding their long-term effects on reducing infectious disease outbreaks in preschool and school contexts.

Together, these studies reinforce the need for prioritising hand hygiene education and interventions as a cost-effective strategy to promote community health and reduce the burden of infectious diseases. Future research should focus on tailoring these interventions to diverse child education settings to maximise their impact and sustainability.

Hand hygiene interventions, particularly handwashing, play a crucial role in mitigating infectious disease outbreaks in children. Promoting hand hygiene in preschools and schools is essential for protecting children’s health and minimising disruptions to education (5–7). Further research is needed to tailor and implement sustainable hand hygiene programs in diverse preschool settings.

### III. Relevance

The success of the Five Moments for Hand Hygiene program in hospital settings demonstrates the transformative impact of structured hand hygiene initiatives on reducing infection rates. By emphasising key moments for hand hygiene: before patient contact, before aseptic tasks, after exposure to body fluids, after patient contact, and after contact with patient surroundings, this program has significantly reduced healthcare-associated infections worldwide (4). Its effectiveness inspires a model that could be adapted for school environments to improve hand hygiene practices and reduce infectious disease transmission. Implementing a similar structured approach to hand hygiene in preschools and schools, focusing on critical moments such as before and after entering the education space, before handling food, after using the restroom, or before and after playing with shared toys, could help mitigate the occurrence of respiratory outbreaks and promote overall health and well-being among this population. However, at present, no such initiative is as universally adopted.

For individual health, reducing the incidence of respiratory and gastrointestinal infections in preschool- and school-aged children, their immediate health may be safeguarded whilst promoting uninterrupted learning and development. From a public health perspective, effective interventions in educational environments can limit disease outbreaks, reduce transmission within communities, and alleviate the burden on families and caregivers. In healthcare, improved hand hygiene leads to fewer infections, decreasing hospital visits, treatment costs, and resource strain. Policy-wise, the findings may be used to guide the development of local or national guidelines for structured hand hygiene programs in early education settings, inspired by successful models like the WHO’s Five Moments for Hand Hygiene (4). Research implications include addressing gaps in understanding the relationship between hand hygiene and infection rates in young children, exploring intervention designs, and evaluating long-term effectiveness to inform broader health strategies.

### IV. Rationale

Paediatric infectious disease outbreaks are a major concern of the general public impacting school absenteeism and parental productivity inferring educational and economic implications. Concerningly, these outbreaks remain apparent in lower-aged groups, especially in preschool and school-aged children. Although contact precautions, annual immunisations, school closures, and isolation methods are ways to mitigate infectious disease transmission rates, the relationship between hand hygiene practices of children is notably less documented.

### V. Justification

The need for this review is justified given the significant health implications and current limitations of the evidence base. Children are highly vulnerable to infectious diseases due to their close interactions and underdeveloped hygiene practices, making this a critical population for targeted interventions. While the effectiveness and practicality of hand hygiene in healthcare settings are well-documented, there is a notable gap in evidence regarding its specific impact on respiratory and gastrointestinal disease outbreaks in child education environments. Addressing this gap is essential to inform tailored strategies that protect children’s health, reduce absenteeism, and mitigate broader public health risks. Moreover, the review’s focus on structured hand hygiene programs has the potential to guide policy development and improve practices in preschools and schools, an area currently underrepresented in the literature. By addressing these limitations, the review will provide valuable insights to guide the development and implementation of effective hand hygiene interventions tailored to these educational environments.

### VI. Specification

PICO components: In paediatric populations, how do hand hygiene practices impact the prevalence of paediatric infectious disease outbreaks?

## 2. Methods

### I. Search strategy Databases

A literature search will be conducted in the electronic databases MEDLINE Complete, Embase, and PubMed as they have proven worth producing credible literature covering a wide range of scientific, medical, and healthcare disciplines. In addition, the Education Resources Information Center (ERIC) electronic database will permit literature from the educational perspective of disease burden, thus adequate for the proposed research question.

#### Key search terms

The keywords or MeSH (Medical Subject Headings) to be used in the search are ‘respiratory tract infection, common cold, influenza, Coronavirus, respiratory syncytial virus, viral gastroenteritis, salmonella, campylobacter, Escherichia coli, shigella, staphylococcus aureus’. Population-defining terms used will be ‘student, school, preschool, daycare, child, children, or infants’. For the intervention, proximity operators will be utilised to within seven words between ‘hand’ and ‘wash, disinfect, sanitise, clean, or hygiene’.

To generate a valid script, the terms needed to be contemplated to ensure the results were relevant whilst not being over-limited. In addition, terminology variation was considered to permit international applications. To achieve this, truncation methods and Boolean operators were applied as a strategic means to produce relevant results for possible inclusion.

This resulted in the following action:

> *(MeSH: respiratory tract infection OR common cold OR influenza OR Coronavirus OR respiratory syncytial virus OR Viral gastroenteritis OR salmonella OR campylobacter OR Escherichia coli OR shigella OR staphylococcus aureus)*
>
> *AND*
>
> *(student OR school OR preschool OR daycare OR child OR children OR infants)*
>
> *AND*
>
> *(hand adj7 (wash* OR disinfect* OR saniti* OR clean* OR hygiene))*

### II. Selection criteria

For the systematic review, the Cochrane methodology will be applied to determine the effect and relationship between hand hygiene practices of children and paediatric respiratory infectious disease prevalence. Each reviewer independently will perform a literature search and screen articles for inclusion, blinded to the other’s decisions.

Following the preliminary search, article duplicates will be identified and removed. The screening process will use the search engine limiters for literature published in the English language and published since 1st January 2014 to ensure currency. Additional screening is limited to academic journals to ensure quality. The first and second pass exclusion will be conducted by assessing the article’s title and then abstract for comparability eliminating any non-comparable intervention or population characteristic. Third-pass exclusion will occur by assessing the full article ensuring that the focused variables are identifiable. Articles are considered for inclusion provided they were of appropriate design, remained after the screening, and no exclusion criteria were identified.

#### Additional limits

Limiters for literature published in the English language and published since 1st January 2014 to ensure currency.

#### Study selection

Data extraction will be done by following the Preferred Reporting Items for Systematic Reviews and Meta-Analyses (PRISMA) guidelines (5, 8).

The reviewers will independently compile inclusion lists. Once completed, the two lists will be compared, and articles selected by both authors will be automatically included in the review. In contrast, articles selected by only one author will undergo further discussion using the Critical Appraisal Skills Programme methodology to reach a consensus on their inclusion. Any articles not selected by either author are excluded from the review. This approach ensures objectivity and rigour in the selection process.

### III. Quality assessment

The Critical Appraisal Skills Programme will be utilised to ensure quality and transparency of selection or exclusion from the study.

### IV. Data extraction

For each included study, data will be extracted into a standardised electronic spreadsheet, including author, year of publication, country, study design, sample size, population characteristics (e.g., age, setting), intervention details (e.g., type of hand hygiene, frequency), outcome measures (e.g., infectious disease incidence, absenteeism), and comment on the major findings.

### V. Data synthesis

Given that the review will involve subgroup analysis of studies focusing on school and preschool-aged children, the synthesis will primarily be a narrative synthesis with a focus on subgroup differences.

#### Subgroup Analysis

The review will perform a subgroup analysis to examine the impact of hand hygiene practices on infectious disease outcomes specifically in preschool (ages 2-4 years) and school populations (ages 5-18 years). This will allow for the identification of patterns or differences in the effectiveness of hand hygiene interventions across various educational settings. Studies will be grouped based on factors such as the type of hand hygiene practice (e.g., handwashing vs. sanitising), the frequency of intervention, and the specific infectious diseases targeted (e.g., respiratory infections, influenza, viral gastroenteritis).

#### Qualitative Synthesis

A narrative synthesis will summarise the findings within these subgroups. This will include a detailed analysis of how hand hygiene practices are implemented and their impact on reducing the incidence of infectious disease outbreaks. The synthesis will consider contextual factors such as the level of adherence to hand hygiene practices, the involvement of staff, and how these interventions might differ based on the type of infectious pathogen being targeted.

## 3. Process

### I. Resources required Relevant Expertise

Expertise in public health, paediatric infectious diseases, epidemiology, and systematic review methodology is necessary.

Familiarity with Cochrane methodology, PRISMA guidelines, and critical appraisal techniques is essential.

#### Computing Facilities

Access to computers or workstations capable of supporting bibliographic and statistical software. Stable internet connection for database searches and accessing academic journals.

#### Research Databases

Access to MEDLINE Complete, Embase, PubMed, and Education Resources Information Center (ERIC) for comprehensive literature retrieval.

#### Bibliographic Software

Software such as EndNote or Mendeley to manage citations and references systematically. For the review, EndNote will be the preferred software utilised.

#### Statistical Software

Software like R, RStudio, or Stata to analyse data if quantitative synthesis is performed.

### II. Dissemination of findings

#### Target Audience

Public health professionals, paediatric healthcare providers, educators, and policymakers that are involved in early childhood education and infection control.

Researchers in the fields of infectious diseases, public health, and paediatric health.

#### Publication Type

The findings will likely be published in peer-reviewed public health, epidemiology, or paediatric healthcare journals, as well as educational journals focusing on infection prevention and control. The intention is for a two-part series, preschool ages (2-4 years) will be reported on in Part 1, while school-aged (5-18 years) children will be included in Part 2 of the publication.

A possible report or policy brief targeting policymakers and education administrators.

#### Communication Media

Academic journal articles and possible conference presentations, webinars or workshops aimed at both healthcare professionals and educators.

Use of digital platforms (e.g., social media, academic networking sites) to promote awareness among a broader audience, including parents and the general public.

## 4. Conclusion

This systematic review protocol aims to fill a critical gap in the literature by evaluating the impact of hand hygiene practices on the reduction of infectious disease outbreaks in educational settings. The proposed review will provide evidence to support the adaptation of successful hospital-based hygiene models for use in preschools and schools, with the goal of improving health and reducing absenteeism among children. Given the potential for widespread public health benefits, the review findings will be crucial for guiding future policy decisions and the development of targeted interventions to protect vulnerable paediatric populations. By addressing this gap in research, the review will contribute to advancing public health strategies and promoting healthier, more resilient educational environments.

## Data Availability

All data produced in the present study are available upon reasonable request to the authors

## Declarations

### Abbreviations

Not applicable

### Ethics Declaration

This study did not require ethical approval as it involved a retrospective analysis of publicly available and anonymised information, with no direct involvement of human subjects.

### Human Ethics

Ethics approval is not required by the Western Sydney University Human Research Ethics Committee (HREC) for this review.

### Consent for publication

The authors consent to the publication of this article.

### Availability of data and materials

Available at request from the corresponding author.

### Authors’ contributions

The authors equally contributed to the conception, design, analysis, and drafting of the manuscript.

## Acknowledgements

Not applicable

## Registration

The systematic review protocol was prospectively submitted to PROSPERO with registration granted on the 10th of December 2024 (CRD42024620293) (9).

## Clinical trial number

not applicable

